# Hearing loss after exposure to vincristine and platinum-based chemotherapy among childhood cancer survivors

**DOI:** 10.1101/2023.03.02.23286688

**Authors:** Sven Strebel, Luzius Mader, Philippa Jörger, Nicolas Waespe, Seraina Uhlmann, Nicolas von der Weid, Marc Ansari, Claudia E. Kuehni

**Affiliations:** Institute of Social and Preventive Medicine, University of Bern, Bern, Switzerland; CANSEARCH research platform in pediatric oncology and hematology, Department of Pediatrics, Gynecology and Obstetrics, University of Geneva, Geneva, Switzerland; Graduate School for Health Sciences, University of Bern, Bern, Switzerland; Division of Pediatric Hematology/Oncology, Department of Pediatrics, Inselspital, Bern University Hospital, University of Bern, Bern, Switzerland; Department of Pediatric Oncology and Hematology, University Children’s Hospital Basel (UKBB), University of Basel, Basel, Switzerland; Department of Women, Child and Adolescent, Division of Pediatric Oncology and Hematology, Geneva University Hospital, University of Geneva, Geneva, Switzerland

**Keywords:** vinca alkaloids, vincristine, hearing loss, ototoxicity, platinum compounds, cranial radiotherapy, childhood cancer survivors

## Abstract

**Background:** Vincristine poses a suspected additional risk factor for hearing loss among childhood cancer survivors (CCS) treated with platinum-based chemotherapy, yet evidence is scarce since no study reports vincristine doses. We examined the association of vincristine with hearing loss in a national cohort of CCS.

**Methods:** We included CCS registered in the Swiss Childhood Cancer Registry treated at age ≤ 18 years with platinum-based chemotherapy between 1990–2014. All participants in our retrospective cohort study had audiogram and treatment data from medical records. We identified CCS exposed to vincristine and calculated the total cumulative dose. We defined clinically relevant hearing loss as grade ≥ 2 using the International Society of Pediatric Oncology Boston Ototoxicity Scale at latest follow-up.

**Results:** Our study population included 270 CCS (43% female; median age at cancer diagnosis 6.8 years; interquartile range [IQR]: 2.1–11.7 years) with median age at audiogram 13.5 years (IQR: 9.3–17.0 years). Vincristine exposure was associated with an increased risk of hearing loss in the multivariable logistic regression analysis (odds ratio [OR] 4.8; 95% confidence interval [CI]: 1.8–12.9). We found no evidence of dose-response relationship (OR 1.0; 95% CI: 0.97–1.04) or effect modification from vincristine from other ototoxic treatments, such as type of platinum agent, cranial radiotherapy, and hematopoietic stem cell transplantation.

**Conclusion:** Vincristine is associated with a higher risk of hearing loss in CCS treated with platinum-based chemotherapy. We suggest future studies investigate the underlying mechanism and causality among CCS without exposure to other ototoxic cancer treatments.

## 1 Introduction

Hearing loss is a side effect of platinum-based chemotherapy among children.[1] It is usually irreversible and impairs neurocognitive functioning of childhood cancer survivors (CCS).[2, 3] Several studies identified risk factors for platinum-induced hearing loss such as age at diagnosis, type of platinum agent, total cumulative dose of platinum, concomitant cranial radiotherapy (CRT), and hematopoietic stem cell transplantation (HSCT).[4-7] However, such factors insufficiently explain interindividual variation of platinum-induced hearing loss, possibly involving additional factors, such as vincristine.[4, 8, 9] Vincristine is known as neurotoxic and reported to cause central neuropathy with paralysis of the auditory nerve (cranial nerve VIII).[10, 11] In addition, vincristine-induced toxicity possibly affects the medial olivocochlear bundle and—to a lesser extent—the outer hair cells.[12]

Only two studies examined the role of vincristine in platinum-induced hearing loss, highlighting the need for further evidence.[4, 9] Both included vincristine as a co-variable in their analysis but did not further investigate the effect of the total cumulative dose of vincristine and whether there was effect modification between vincristine and other ototoxic cancer treatments. Therefore, we analyzed data from a nationwide cohort of CCS with treatment and audiogram data from medical records to 1) quantify the effect of vincristine on platinum-induced hearing loss; 2) test for interactions between vincristine and other ototoxic cancer treatments; and 3) determine a possible dose-response relationship of vincristine-induced ototoxicity.

## 2. Methods

### 2.1. Study population

We analyzed data from CCS treated with platinum-based chemotherapy between 1990– 2014 in the nine specialized pediatric oncology clinics in Switzerland.[6] We included CCS 1) registered in the national and population-based Swiss Childhood Cancer Registry (SCCR),[13] 2) treated for cancer at age ≤ 18 years, 3) with no evidence of hearing loss before start of cancer treatment in their medical records, and 4) available audiogram after completing platinum-based chemotherapy.[6] Further details about identifying eligible CCS and data collection are published elsewhere.[6, 14] The Ethics Committee of the Canton of Bern approved the SCCR and the Swiss Childhood Cancer Survivor Study (KEK-BE: 166/2014; 2021-01462).[15]

### 2.2. Study procedure

We collected clinical information on cancer diagnosis and treatment from medical records and the SCCR in 2015.[6, 14] We extracted clinical variables, including sex (female, male); cancer diagnosis according to the International Classification of Childhood Cancer (3^rd^ edition)[16]; age at cancer diagnosis (< 5, 5–9, and 10–18 years); period of cancer diagnosis (1990–1995, 1996–2001, 2002–2007, and 2008–2014); type and total cumulative dose (mg/m^2^) of platinum agents (cisplatin, carboplatin); CRT (yes, no); and HSCT (yes, no).

During initial data collection in 2015, no treatment information about vincristine exposure was collected. We went through all medical records and treatment protocols again in 2022 and calculated vincristine exposure and total cumulative vincristine dose (mg/m^2^) as treated or— when unavailable—based on treatment protocol.

### 2.3. Measurement of hearing loss

We determined hearing loss at latest follow-up after completion of platinum-based chemotherapy. We evaluated audiograms with the classification system of the International Society of Pediatric Oncology (SIOP) Boston Ototoxicity Scale.[17] Based on the approach of previous studies, we defined clinically relevant hearing loss (yes, no) as SIOP-Boston grade ≥ 2.[4, 6] In cases of asymmetric hearing, we took the most affected ear as the reference for grading. Further details on the evaluation of the audiograms were previously published.[6]

### 2.4. Statistical analysis

First, we compared the prevalence of clinically relevant hearing loss overall and stratified by type of platinum-based chemotherapy between CCS with and without vincristine exposure. We fitted univariable logistic regression models including sex, age at audiogram, age at cancer diagnosis, period of cancer diagnosis, type of platinum-based chemotherapy, exposure to vincristine, concomitant CRT, and HSCT as exposures and clinically relevant hearing loss (yes, no) as outcome.[6] The multivariable regressions included a priori sex, age at most recent audiogram, age at cancer diagnosis, and all other clinical characteristics previously associated with hearing loss at *P* < 0.05 in the univariable models. To examine effect modification of other ototoxic treatments, we included interaction terms between vincristine exposure and type of platinum-based chemotherapy, CRT, and HSCT.[4, 18, 19] We also included the total cumulative dose of cisplatin, carboplatin, and vincristine as a continuous variable in a sub-analysis, which included only CCS who received vincristine, to determine a possible dose-response relationship. We excluded survivors with missing values for the cumulative dose of vincristine or platinum-based chemotherapy from the regression analysis. We calculated global *P*-values using likelihood-ratio tests (LRT).

We performed a sensitivity analysis to control for additional ototoxic factors, such as brain surgeries or cerebrospinal fluid shunts, associated with central nervous system (CNS) tumors diagnosis.[1, 20] We included CNS tumor diagnosis (yes, no) as a co-variable in our multivariable regression model.

We used Stata version 16.1 (StataCorp LP, Austin, TX, USA) for all analyses.

## 3. Results

### 3.1. Characteristics of study population

We included 270 CCS with a median age at cancer diagnosis of 6.8 years (interquartile range [IQR]: 2.1–11.7 years) in our analysis (Table 1). A detailed flow-diagram of the study population was published elsewhere.[6] Median time from cancer diagnosis to most recent audiogram was 5 years (IQR 2.5–8.1 years). The most common cancer diagnoses were CNS tumors (n = 104; 39%). Over half (n = 145; 54%) received vincristine with a median total cumulative dose of 23 mg/m^2^ (IQR 10.1–40.9 mg/m^2^) (Supplement Figure A.1). CCS previously treated with vincristine were younger at cancer diagnosis, more likely survivors of CNS tumors or neuroblastoma, and received more often carboplatin, CRT, and HSCT than CCS without vincristine exposure.

**TABLE 1.** Demographic and clinical characteristics of study population

|  | Total cohort<br>N=270 |  | No Vincristine<br>n=125 |  | Vincristine<br>n=145 |  | P-value<sup>a</sup> |
| --- | --- | --- | --- | --- | --- | --- | --- |
| Demographic characteristics | n | (%) | n | (%) | n | (%) |  |
| *Sex* |  |  |  |  |  |  | 0.192 |
| Male | 154 | (57) | 66 | (53) | 88 | (61) |  |
| Female | 116 | (43) | 59 | (47) | 57 | (39) |  |
| *Age at most recent audiogram* |  |  |  |  |  |  | 0.066 |
| < 10 years | 81 | (30) | 32 | (26) | 49 | (34) |  |
| 10–15 years | 108 | (40) | 47 | (38) | 61 | (42) |  |
| > 15 years | 81 | (30) | 46 | (37) | 35 | (24) |  |
| Clinical characteristics | n | (%) | n | (%) | n | (%) |  |
| *Age at cancer diagnosis* |  |  |  |  |  |  | <0.001 |
| < 5 years | 112 | (41) | 42 | (34) | 70 | (48) |  |
| 5–9 years | 67 | (25) | 24 | (19) | 43 | (30) |  |
| 10–18 years | 91 | (34) | 59 | (47) | 32 | (22) |  |
| *Period of cancer diagnosis* |  |  |  |  |  |  | 0.454 |
| 1990–1995 | 48 | (18) | 24 | (19) | 24 | (17) |  |
| 1996–2001 | 75 | (28) | 36 | (29) | 39 | (27) |  |
| 2002–2007 | 82 | (30) | 32 | (26) | 50 | (34) |  |
| 2008–2014 | 65 | (24) | 33 | (26) | 32 | (22) |  |
| *Diagnosis (ICCC-3)* |  |  |  |  |  |  | <0.001 |
| III CNS and miscellaneous intracranial and intraspinal neoplasms | 104 | (39) | 3 | (2) | 101 | (70) |  |
| IV Neuroblastoma and other peripheral nervous cell tumors | 39 | (14) | 13 | (10) | 26 | (18) |  |
| V Retinoblastoma | 5 | (2) | 2 | (2) | 3 | (2) |  |
| VI Renal tumors | 6 | (2) | 3 | (2) | 3 | (2) |  |
| VII Hepatic tumors | 15 | (6) | 14 | (11) | 1 | (1) |  |
| VIII Malignant Bone tumors | 62 | (23) | 62 | (50) | 0 | (0) |  |
| IX Soft tissue and other extraosseous sarcoma | 12 | (4) | 1 | (1) | 11 | (8) |  |
| X Germ cell tumors, trophoblastic tumors, and neoplasms of gonads | 27 | (10) | 27 | (22) | 0 | (0) |  |
| *Treatments<sup>b</sup>* |  |  |  |  |  |  |  |
| *Platinum agent* |  |  |  |  |  |  | <0.001 |
| Cisplatin | 151 | (56) | 98 | (78) | 53 | (37) |  |
| Carboplatin | 62 | (23) | 17 | (14) | 45 | (31) |  |
| Cisplatin and Carboplatin | 57 | (21) | 10 | (8) | 47 | (32) |  |
| *Cumulative cisplatin dose categories* |  |  |  |  |  |  | <0.001 |
| No cisplatin | 62 | (23) | 17 | (14) | 45 | (31) |  |
| ≤ 300 mg/m<sup>2</sup> | 51 | (19) | 11 | (9) | 40 | (28) |  |
| 301–450 mg/m<sup>2</sup> | 71 | (26) | 38 | (30) | 33 | (23) |  |
| > 450 mg/m<sup>2</sup> | 86 | (32) | 59 | (47) | 27 | (19) |  |
| *Cumulative carboplatin dose categories* |  |  |  |  |  |  | <0.001 |
| No carboplatin | 151 | (56) | 98 | (78) | 53 | (37) |  |
| < 1500 mg/m<sup>2</sup> | 29 | (11) | 3 | (2) | 26 | (18) |  |
| 1500–3000 mg/m<sup>2</sup> | 53 | (20) | 14 | (11) | 39 | (27) |  |
| > 3000 mg/m<sup>2</sup> | 32 | (12) | 6 | (5) | 26 | (18) |  |
| Missing | 5 | (2) | 4 | (3) | 1 | (1) |  |
| *Cranial radiation* |  |  |  |  |  |  | <0.001 |
| No | 172 | (64) | 115 | (92) | 57 | (39) |  |
| Yes | 98 | (36) | 10 | (8) | 88 | (61) |  |
| *HSCT* |  |  |  |  |  |  | 0.004 |
| No | 244 | (90) | 120 | (96) | 124 | (86) |  |
| Yes | 26 | (10) | 5 | (4) | 21 | (14) |  |

Abbreviations: CNS, central nervous system; ICCC-3, International Classification of Childhood Cancer–Third Edition; HSCT, hematopoietic stem cell transplantation.
<sup>a</sup>P-values calculated from chi-square statistics comparing childhood cancer survivors with and without vincristine exposure.
<sup>b</sup>Each subject could have had more than one treatment modality.

### 3.2. Role of vincristine in platinum-induced hearing loss

Of the included CCS treated with vincristine, 49% (95% CI: 41-57) had hearing loss compared with 22% (95% CI: 15-30) of CCS not treated with vincristine (*p* < 0.001) (Figure 1). Stratified by type of platinum-based chemotherapy, we saw the largest difference in prevalence of hearing loss between CCS with and without vincristine exposure among those treated with only cisplatin (68% vs. 21%).

**FIGURE 1.**
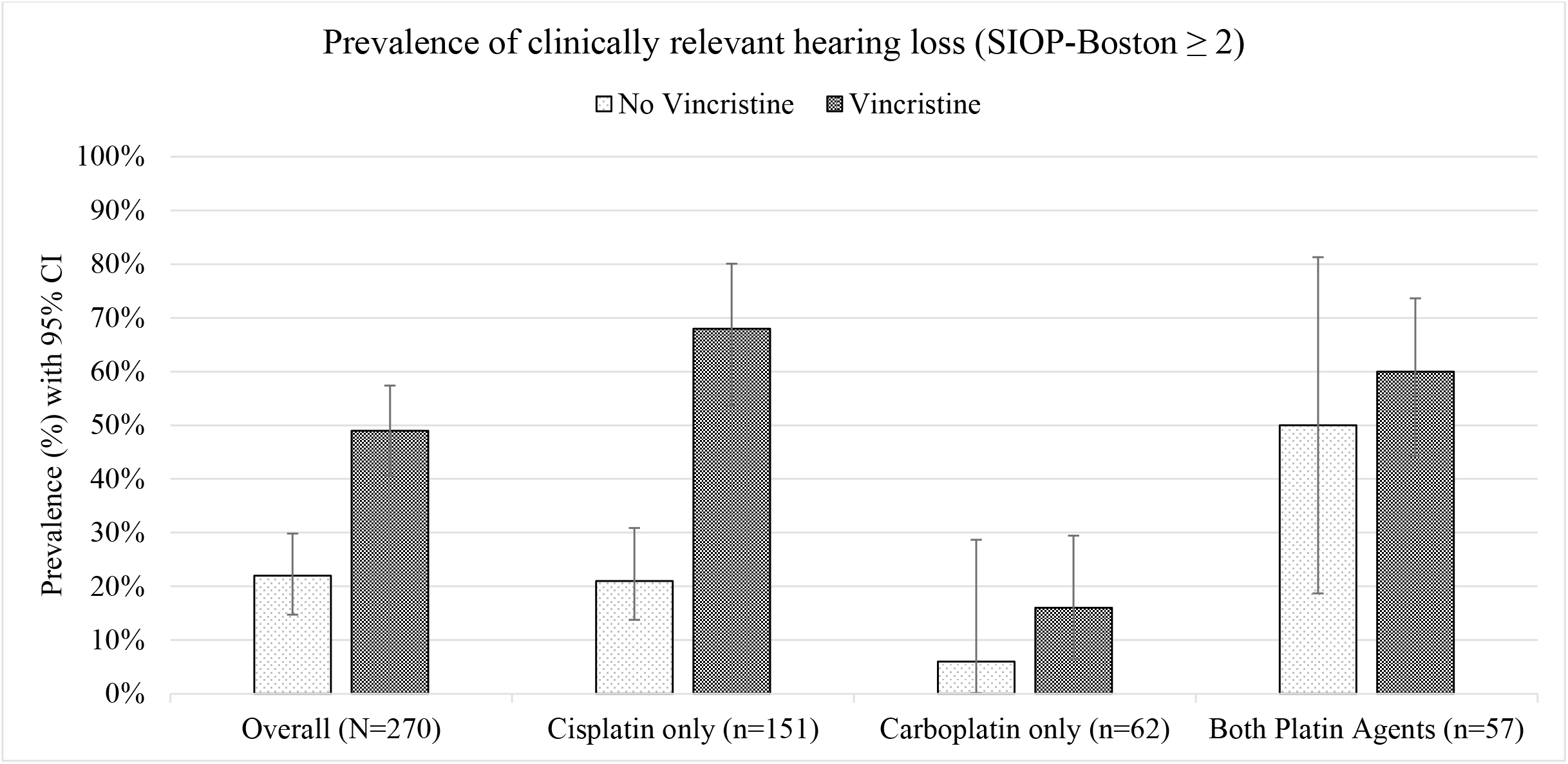
Comparison of prevalence of clinically relevant hearing loss between childhood cancer survivors with and without vincristine exposure (N=270). Prevalence is shown for the overall cohort and stratified by type of platinum-based chemotherapy.

Exposure to vincristine was associated with a higher risk for hearing loss in the univariable (odds ratio [OR] 3.5, 95% CI: 2.0–6.0) and multivariable (OR 4.8, 95% CI: 1.8–12.9) logistic regression analyses (Table 2). Using interaction terms, we found no evidence for effect modification between exposure to vincristine and type of platinum-based chemotherapy, CRT, and HSCT (all *P*>0.05) (Table 3). In a sub-analysis including only CCS with vincristine exposure, we found no evidence for a linear effect of total cumulative vincristine dose on the risk of hearing loss (OR 1.0, 95% CI: 0.97–1.04) (Table 2) (Supplement Figure A.1).

**TABLE 2.**
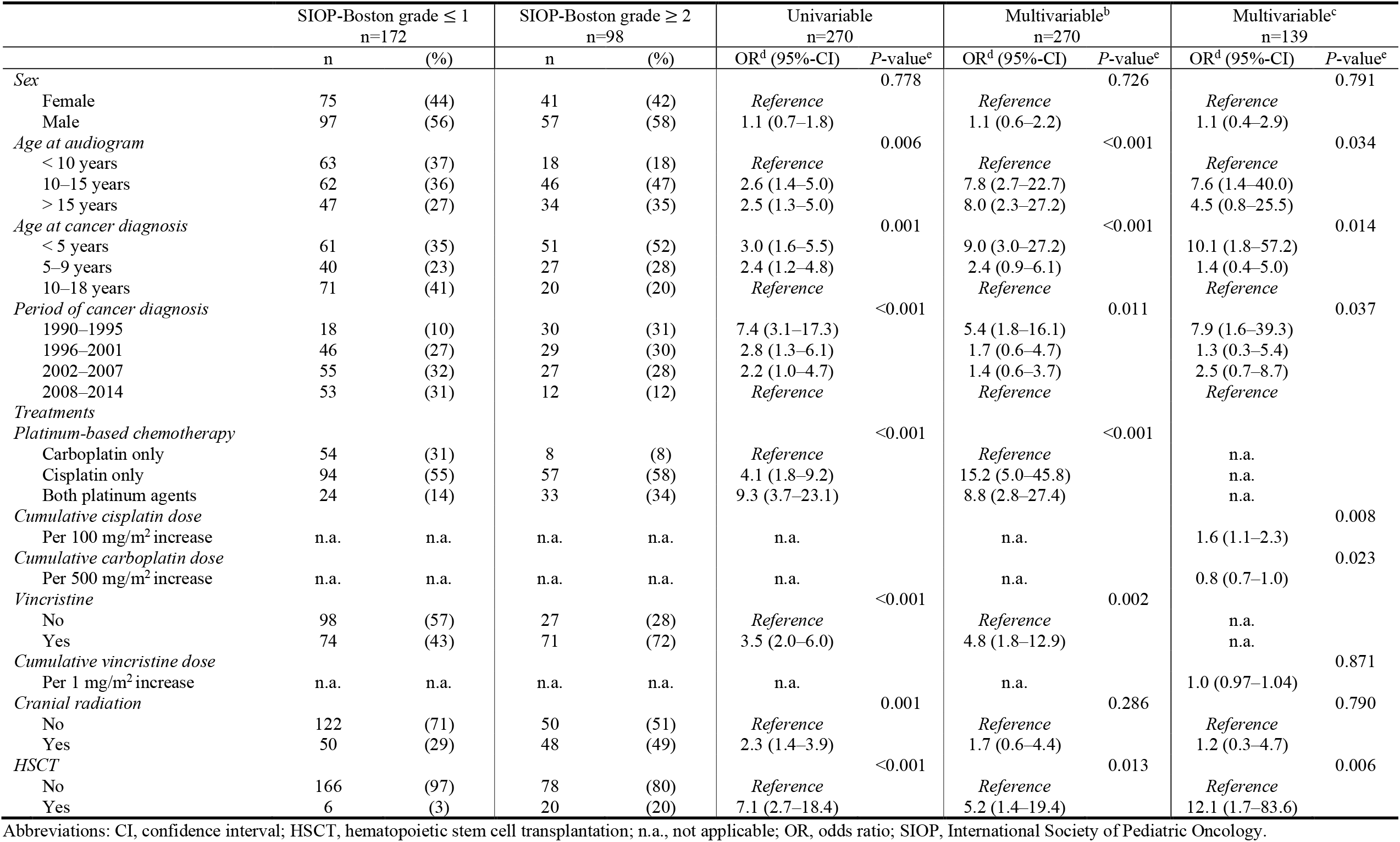
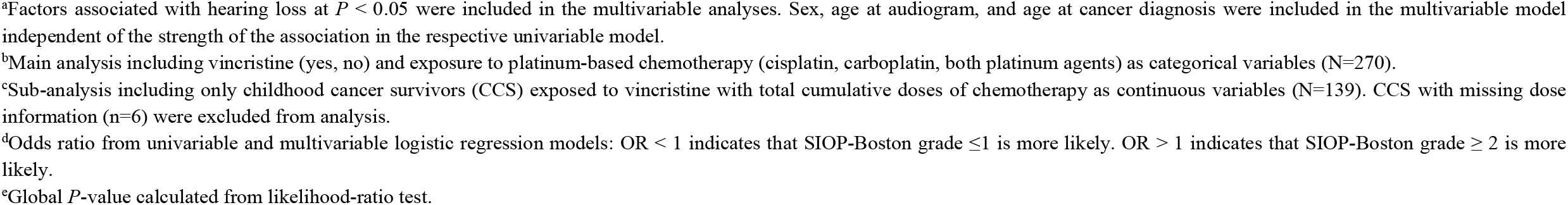
Results from univariable and multivariable^a^ logistic regression analyses

|  | SIOP-Boston grade ≤ 1<br>n=172 |  | SIOP-Boston grade ≥ 2<br>n=98 |  | Univariable<br>n=270 |  | Multivariable<sup>b</sup><br>n=270 |  | Multivariable<sup>c</sup><br>n=139 |  |
| --- | --- | --- | --- | --- | --- | --- | --- | --- | --- | --- |
|  | n | (%) | n | (%) | OR<sup>d</sup> (95%-CI) | P-value<sup>e</sup> | OR<sup>d</sup> (95%-CI) | P-value<sup>e</sup> | OR<sup>d</sup> (95%-CI) | P-value<sup>e</sup> |
| *Sex* |  |  |  |  |  | 0.778 |  | 0.726 |  | 0.791 |
| Female | 75 | (44) | 41 | (42) | Reference |  | Reference |  | Reference |  |
| Male | 97 | (56) | 57 | (58) | 1.1 (0.7–1.8) |  | 1.1 (0.6–2.2) |  | 1.1 (0.4–2.9) |  |
| *Age at audiogram* |  |  |  |  |  | 0.006 |  | <0.001 |  | 0.034 |
| < 10 years | 63 | (37) | 18 | (18) | Reference |  | Reference |  | Reference |  |
| 10–15 years | 62 | (36) | 46 | (47) | 2.6 (1.4–5.0) |  | 7.8 (2.7–22.7) |  | 7.6 (1.4–40.0) |  |
| > 15 years | 47 | (27) | 34 | (35) | 2.5 (1.3–5.0) |  | 8.0 (2.3–27.2) |  | 4.5 (0.8–25.5) |  |
| *Age at cancer diagnosis* |  |  |  |  |  | 0.001 |  | <0.001 |  | 0.014 |
| < 5 years | 61 | (35) | 51 | (52) | 3.0 (1.6–5.5) |  | 9.0 (3.0–27.2) |  | 10.1 (1.8–57.2) |  |
| 5–9 years | 40 | (23) | 27 | (28) | 2.4 (1.2–4.8) |  | 2.4 (0.9–6.1) |  | 1.4 (0.4–5.0) |  |
| 10–18 years | 71 | (41) | 20 | (20) | Reference |  | Reference |  | Reference |  |
| *Period of cancer diagnosis* |  |  |  |  |  | <0.001 |  | 0.011 |  | 0.037 |
| 1990–1995 | 18 | (10) | 30 | (31) | 7.4 (3.1–17.3) |  | 5.4 (1.8–16.1) |  | 7.9 (1.6–39.3) |  |
| 1996–2001 | 46 | (27) | 29 | (30) | 2.8 (1.3–6.1) |  | 1.7 (0.6–4.7) |  | 1.3 (0.3–5.4) |  |
| 2002–2007 | 55 | (32) | 27 | (28) | 2.2 (1.0–4.7) |  | 1.4 (0.6–3.7) |  | 2.5 (0.7–8.7) |  |
| 2008–2014 | 53 | (31) | 12 | (12) | Reference |  | Reference |  | Reference |  |
| *Treatments* |  |  |  |  |  |  |  |  |  |  |
| *Platinum-based chemotherapy* |  |  |  |  |  | <0.001 |  | <0.001 |  |  |
| Carboplatin only | 54 | (31) | 8 | (8) | Reference |  | Reference |  | n.a. |  |
| Cisplatin only | 94 | (55) | 57 | (58) | 4.1 (1.8–9.2) |  | 15.2 (5.0–45.8) |  | n.a. |  |
| Both platinum agents | 24 | (14) | 33 | (34) | 9.3 (3.7–23.1) |  | 8.8 (2.8–27.4) |  | n.a. |  |
| *Cumulative cisplatin dose* |  |  |  |  |  |  |  |  |  | 0.008 |
| Per 100 mg/m<sup>2</sup> increase | n.a. | n.a. | n.a. | n.a. | n.a. |  | n.a. |  | 1.6 (1.1–2.3) |  |
| *Cumulative carboplatin dose* |  |  |  |  |  |  |  |  |  | 0.023 |
| Per 500 mg/m<sup>2</sup> increase | n.a. | n.a. | n.a. | n.a. | n.a. |  | n.a. |  | 0.8 (0.7–1.0) |  |
| *Vincristine* |  |  |  |  |  | <0.001 |  | 0.002 |  |  |
| No | 98 | (57) | 27 | (28) | Reference |  | Reference |  | n.a. |  |
| Yes | 74 | (43) | 71 | (72) | 3.5 (2.0–6.0) |  | 4.8 (1.8–12.9) |  | n.a. |  |
| *Cumulative vincristine dose* |  |  |  |  |  |  |  |  |  | 0.871 |
| Per 1 mg/m<sup>2</sup> increase | n.a. | n.a. | n.a. | n.a. | n.a. |  | n.a. |  | 1.0 (0.97–1.04) |  |
| *Cranial radiation* |  |  |  |  |  | 0.001 |  | 0.286 |  | 0.790 |
| No | 122 | (71) | 50 | (51) | Reference |  | Reference |  | Reference |  |
| Yes | 50 | (29) | 48 | (49) | 2.3 (1.4–3.9) |  | 1.7 (0.6–4.4) |  | 1.2 (0.3–4.7) |  |
| *HSCT* |  |  |  |  |  | <0.001 |  | 0.013 |  | 0.006 |
| No | 166 | (97) | 78 | (80) | Reference |  | Reference |  | Reference |  |
| Yes | 6 | (3) | 20 | (20) | 7.1 (2.7–18.4) |  | 5.2 (1.4–19.4) |  | 12.1 (1.7–83.6) |  |

Abbreviations: CI, confidence interval; HSCT, hematopoietic stem cell transplantation; n.a., not applicable; OR, odds ratio; SIOP, International Society of Pediatric Oncology.
<sup>a</sup>Factors associated with hearing loss at $P < 0.05$ were included in the multivariable analyses. Sex, age at audiogram, and age at cancer diagnosis were included in the multivariable model independent of the strength of the association in the respective univariable model.
<sup>b</sup>Main analysis including vincristine (yes, no) and exposure to platinum-based chemotherapy (cisplatin, carboplatin, both platinum agents) as categorical variables (N=270).
<sup>c</sup>Sub-analysis including only childhood cancer survivors (CCS) exposed to vincristine with total cumulative doses of chemotherapy as continuous variables (N=139). CCS with missing dose information (n=6) were excluded from analysis.
<sup>d</sup>Odds ratio from univariable and multivariable logistic regression models: OR < 1 indicates that SIOP-Boston grade $\leq 1$ is more likely. OR > 1 indicates that SIOP-Boston grade $\geq 2$ is more likely.
<sup>e</sup>Global $P$ -value calculated from likelihood-ratio test.

**TABLE 3.** Interactions between vincristine and other ototoxic cancer treatments

|  |  | Multivariable<sup>a</sup> |  |
| --- | --- | --- | --- |
|  |  | OR<sup>b</sup> (95%-CI) | P-value<sup>c</sup> |
| **Model 1** |  |  |  |
| *Test for interaction between platinum-based chemotherapy and vincristine* |  |  | 0.274 |
| Carboplatin |  | Reference |  |
| Cisplatin × vincristine exposure |  | 2.8 (0.2–35.2) |  |
| Both platinum agents × vincristine exposure |  | 0.5 (0.03–11.0) |  |
| **Model 2** |  |  |  |
| *Test for interaction between CRT and vincristine* |  |  | 0.200 |
| No CRT |  | Reference |  |
| CRT × vincristine exposure |  | 0.3 (0.04–2.0) |  |
| **Model 3** |  |  |  |
| *Test for interaction between HSCT and vincristine* |  |  | 0.354 |
| No HSCT |  | Reference |  |
| HSCT × vincristine exposure |  | 4.4 (0.18–105.8) |  |

Abbreviations: CI, confidence interval; CRT, cranial radiotherapy; HSCT, hematopoietic stem cell transplantation; OR, odds ratio.
<sup>a</sup>Each interaction between vincristine and the variables platinum-based chemotherapy, CRT, or HSCT was analyzed in a separate multivariable model (Models 1–3) (N=270). Each model was adjusted for sex, age at audiogram, age at diagnosis, period of diagnosis, other ototoxic cancer treatments (i.e. platinum-based chemotherapy, CRT, or HSCT).
<sup>b</sup>Odds ratio from univariable and multivariable logistic regression models: OR < 1 indicates that SIOP-Boston grade ≤ 1 is more likely. OR > 1 indicates that SIOP-Boston grade ≥ 2 is more likely.
<sup>c</sup>Global P-value calculated from likelihood-ratio test.

In our sensitivity analysis, diagnosis with a CNS tumor (yes, no) was not associated with an increased risk for hearing loss in the multivariable regression (OR 0.8, 95% CI: 0.2–2.7) (Supplement Table A.1). The effect size of vincristine (yes, no) did not change compared with the model without adjustment for CNS tumor diagnosis (OR 5.3, 95% CI: 1.7–17.0 vs. OR 4.8, 95% CI: 1.8–12.9).

## 4. Discussion

In our nationwide cohort study of CCS treated with platinum-based chemotherapy, we found the prevalence of platinum-induced hearing loss higher among CCS with additional vincristine than of CCS without vincristine exposure. Vincristine exposure increased the risk of hearing loss fivefold when adjusting for clinical characteristics. However, we found no evidence of a dose-response relationship of cumulative vincristine dose and the risk for hearing loss, or evidence of effect modification by other ototoxic cancer treatments.

Our study is strengthened from analyzing the ototoxicity of vincristine in a large cohort of CCS in terms of overall exposure and considering the total cumulative vincristine dose and potential interactions with other ototoxic cancer treatments. Further strengths include the well-described cohort with clinical data on hearing function, cancer diagnosis, and platinum-based chemotherapy available. However, we cannot conclude whether vincristine is ototoxic independently or only in combination with platinum agents because all included CCS were treated with platinum-based chemotherapy. Furthermore, we had no data on brain surgery or cerebrospinal fluid shunts among survivors of CNS tumors, which have been associated with an increased risk of hearing loss in other studies.[1, 20, 21] Because 70% of CCS in our cohort who received vincristine were diagnosed with CNS tumors and most patients with CNS tumors were exposed to vincristine (97%), we cannot exclude that part of the association of vincristine with hearing loss as possibly explained by other factors associated with the diagnosis of CNS tumors or other concomitant treatments. However, in our sensitivity analysis, CNS tumor diagnosis was not associated with an increased risk of hearing loss and the effect of vincristine remained unchanged after controlling for CNS tumor diagnosis. We also lack data on use of concomitant ototoxic drugs such as aminoglycoside antibiotics and on prophylactic administration of sodium thiosulfate.[1, 22] However, sodium thiosulfate was only recently been approved in the United States for clinical use; it is not yet approved in Switzerland.[23] Another limitation is the retrospective study design and the use of non-standardized routine data from different clinics.

Ototoxicity of vincristine was reported mainly in case studies of adult patients with cancer and in two recent cohort studies involving CCS.[4, 9, 11, 24-27] Moke and colleagues examined hearing loss in a large multi-center cohort of 1,481 children, adolescents, and young adults treated with cisplatin.[4] They also collected data on vincristine exposure and reported that vincristine (yes, no) increased the risk for cisplatin-induced hearing loss by a factor of four (OR 3.6) at latest follow-up,[4] which is consistent with our estimate (OR 4.8). However, a direct comparison must be made with caution because Moke and colleagues also included adults in their analysis and the regression models differed in terms of selected co-variables. In another cohort study of 368 pediatric patients with cancer, Meijer and colleagues demonstrated vincristine exposure (hazard ratio 2.9) was an important determinant for the cumulative incidence of cisplatin-induced hearing loss.[9] Neither of the two cohort studies included data on the cumulative dose of vincristine.[4, 9]

We did not observe an effect from higher cumulative doses of vincristine on the risk of hearing loss, yet we had no data on administration method, timing of administration with other drugs (particularly platinum agents), pharmacokinetics, drug-drug interactions with azole antifungals, and genetic factors possibly affecting total exposure and toxicity of vincristine.[28-31] Two prospective studies with small sample sizes (N ≤ 23) examined ototoxicity of vincristine and included dose information.[12, 32] Lugassy and colleagues enrolled adult patients with cancer (N=23) in their study and did not observe any deleterious effects from vincristine on hearing among patients with moderate dose exposure (mean total dose of 12 mg).[32] One patient on high-dose vincristine (total dose of 24 mg) developed sensorineural hearing loss during the observation period.[32] Riga and colleagues enrolled pediatric patients diagnosed with leukemia in their study (N=15) who had been treated according to the Berlin-Frankfurt-Muenster-95 (BMF-95) protocol. In their study, exposure to low and moderate doses of vincristine (≤ 1.5 mg/m^2^ per dose) did not result in any abnormal changes (≥ 15 dB) in mean hearing thresholds levels measured by pure tone audiometry at latest follow-up.[12] But they found abnormalities in transient evoked otoacoustic emissions (TEOAE) and a decrease in contralateral suppression after three cycles of vincristine administration, indicating a measurable neurotoxic effect of vincristine on outer hair cells and the medial olivocochlear bundle.[12] However, the study’s short observation period of 22 days does not allow prediction of the reversibility of observed effects or subsequent detection of hearing loss.[12]

## 5. Conclusion

Our results support emerging evidence for an increased risk of platinum-induced hearing loss with concomitant vincristine administration.[4, 9] Interindividual variability in hearing loss after platinum-based chemotherapy is large and not yet fully understood.[8], which emphasizes the importance of identifying additional risk factors for hearing loss among CCS, such as the neurotoxic treatment vincristine. However, we suggest including TEOAE measurements in further studies to better understand the mechanism and clinical course of vincristine-induced neurotoxicity in the medial olivocochlear bundle and its influence on platinum-induced hearing loss. In addition, we suggest investigating causality of vincristine and hearing loss among CCS without platinum-based chemotherapy or CRT to determine whether vincristine causes permanent hearing loss even without established ototoxic cancer treatments and whether genetic factors possibly play a role.

## Data Availability

The data that support the information of this manuscript were accessed on secured servers of the Institute of Social and Preventive Medicine at the University of Bern. Individual-level sensitive data can only be made available for researchers who fulfil the respective legal requirements. All data requests should be communicated to the corresponding author.

## Clinical trial registration

Not applicable.

## Funding source

This study was supported by the Swiss Cancer League (HSR-4951-11-2019), the CANSEARCH Foundation (https://cansearch.ch/), and the European Union’s Seventh

Framework Program for research, technological development and demonstration under grant agreement no. 602030. The EU Commission takes no responsibility for any use made of the information set out. Further funding comes from Swiss Cancer Research (KLS/KFS-4825-01-2019, KFS-5027-02-2020) and Kinderkrebshilfe Schweiz (https://www.kinderkrebshilfe.ch/de).

## Declaration of competing interest

The authors declare that they have no known competing financial interests or personal relationships that could have appeared to influence the work reported in this paper.

## Acknowledgments

We thank the study team of the Childhood Cancer Research Group, Institute of Social and Preventive Medicine, University of Bern, and the Swiss Childhood Cancer Survivor Study, the data managers of the Swiss Pediatric Oncology Group, the team of the Swiss Childhood Cancer Registry, and our collaborators of the PanCareLIFE project. We further thank the research group of the CANSEARCH research platform in pediatric oncology and hematology of the University of Geneva for their inputs and support. We thank Kristin Bivens from the editorial service of the Institute of Social and Preventive Medicine at the University of Bern for her editorial suggestions.

## Notes

### Competing Interest Statement

The authors have declared no competing interest.

### Author Declarations

The Ethics Committee of the Canton of Bern approved the SCCR and the Swiss Childhood Cancer Survivor Study (KEK-BE: 166/2014; 2021-01462).

