## Supplementary information for "Hearing loss after exposure to vincristine and platinum-based chemotherapy among childhood cancer survivors"

### Supplementary data

**SUPPLEMENTARY TABLE A.1** Sensitivity analysis including CNS tumor diagnosis into the regression analysis

| | SIOP-Boston grade ≤ 1 n=172 | | SIOP-Boston grade ≥ 2 n=98 | | Univariable | | Multivariable[a] | |
|---|---|---|---|---|---|---|---|---|
| | n | (%) | n | (%) | OR[b] (95%-CI) | *P*-value[c] | OR[b] (95%-CI) | *P*-value[c] |
| *Sex* | | | | | | 0.778 | | 0.728 |
| Female | 75 | (44) | 41 | (42) | *Reference* | | *Reference* | |
| Male | 97 | (56) | 57 | (58) | 1.1 (0.7-1.8) | | 1.1 (0.6-2.2) | |
| *Age at audiogram* | | | | | | 0.006 | | <0.001 |
| < 10 years | 63 | (37) | 18 | (18) | *Reference* | | *Reference* | |
| 10-15 years | 62 | (36) | 46 | (47) | 2.6 (1.4-5.0) | | 8.0 (2.7-23.3) | |
| > 15 years | 47 | (27) | 34 | (35) | 2.5 (1.3-5.0) | | 8.0 (2.3-27.6) | |
| *Age at cancer diagnosis* | | | | | | 0.001 | | <0.001 |
| < 5 years | 61 | (35) | 51 | (52) | 3.0 (1.6-5.5) | | 9.0 (3.0-27.1) | |
| 5-9 years | 40 | (23) | 27 | (28) | 2.4 (1.2-4.8) | | 2.4 (0.9-6.1) | |
| 10-18 years | 71 | (41) | 20 | (20) | *Reference* | | *Reference* | |
| *Period of cancer diagnosis* | | | | | | <0.001 | | 0.011 |
| 1990-1995 | 18 | (10) | 30 | (31) | 7.4 (3.1-17.3) | | 5.4 (1.8-16.1) | |
| 1996-2001 | 46 | (27) | 29 | (30) | 2.8 (1.3-6.1) | | 1.7 (0.6-4.7) | |
| 2002-2007 | 55 | (32) | 27 | (28) | 2.2 (1.0-4.7) | | 1.4 (0.6-3.7) | |
| 2008-2014 | 53 | (31) | 12 | (12) | *Reference* | | *Reference* | |
| *CNS tumor diagnosis* | | | | | | 0.004 | | 0.711 |
| No | 117 | (68) | 49 | (50) | *Reference* | | *Reference* | |
| Yes | 55 | (32) | 49 | (50) | 2.1 (1.3-3.5) | | 0.8 (0.2-2.7) | |
| *Treatments* | | | | | | | | |
| *Platinum-based chemotherapy* | | | | | | <0.001 | | <0.001 |
| Carboplatin only | 54 | (31) | 8 | (8) | *Reference* | | *Reference* | |
| Cisplatin only | 94 | (55) | 57 | (58) | 4.1 (1.8-9.2) | | 15.7 (5.1-48.3) | |
| Both platinum agents | 24 | (14) | 33 | (34) | 9.3 (3.7-23.1) | | 9.2 (2.8-30.0) | |
| *Vincristine* | | | | | | <0.001 | | 0.004 |
| No | 98 | (57) | 27 | (28) | *Reference* | | *Reference* | |
| Yes | 74 | (43) | 71 | (72) | 3.5 (2.0-6.0) | | 5.3 (1.7-17.0) | |
| *Cranial radiation* | | | | | | 0.001 | | 0.265 |
| No | 122 | (71) | 50 | (51) | *Reference* | | *Reference* | |
| Yes | 50 | (29) | 48 | (49) | 2.3 (1.4-3.9) | | 1.9 (0.6-5.6) | |
| *HSCT* | | | | | | <0.001 | | 0.021 |
| No | 166 | (97) | 78 | (80) | *Reference* | | *Reference* | |
| Yes | 6 | (3) | 20 | (20) | 7.1 (2.7-18.4) | | 4.9 (1.3-19.0) | |

Abbreviations: CI, confidence interval; CNS, central nervous system; HSCT, hematopoietic stem cell transplantation, OR, odds ratio; SIOP, International Society of Pediatric Oncology.

[a]Factors associated with hearing loss at $P < 0.05$ were included in the multivariable analyses. Sex, age at audiogram, and age at cancer diagnosis were included in the multivariable model independent of the strength of the association in the respective univariable model.

[b]Odds ratio from univariable and multivariable logistic regression models: OR < 1 indicates that SIOP-Boston grade ≤1 is more likely. OR > 1 indicates that SIOP-Boston grade ≥ 2 is more likely.

non peer-reviewed

[c]Global *P*-value calculated from likelihood-ratio test.

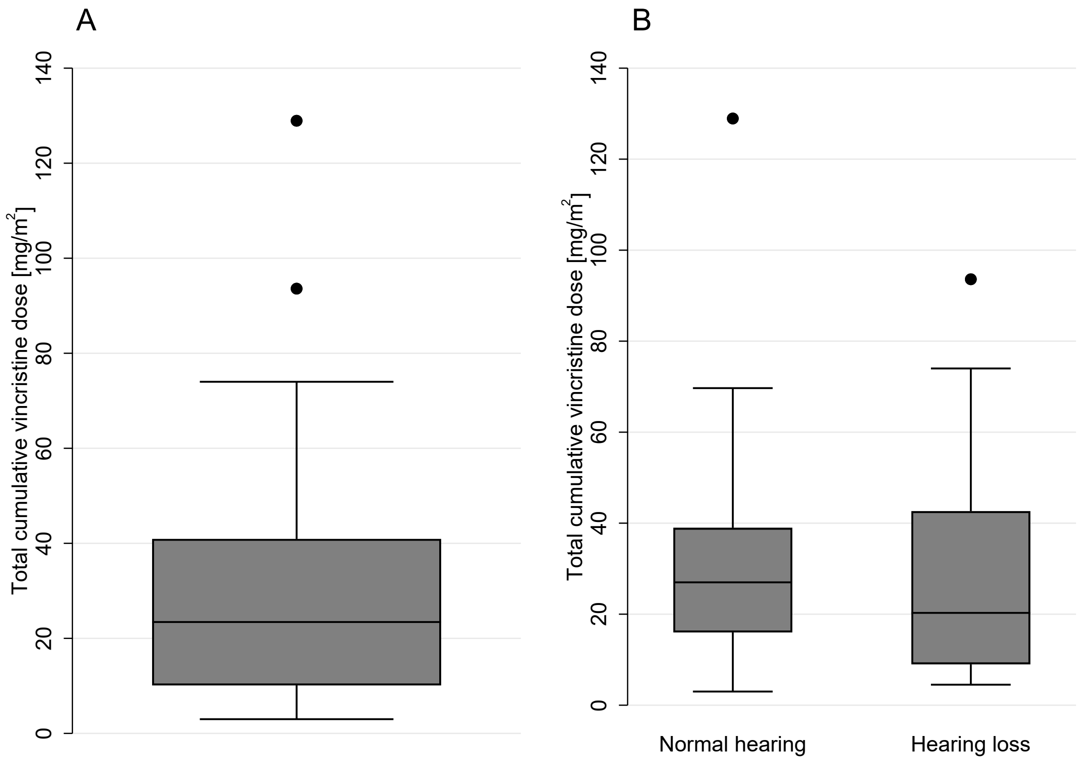


**SUPPLEMENTARY FIGURE A.1** Distribution of the calculated total cumulative vincristine dose overall [A] and stratified by hearing function [B] (n=140, excluding 5 survivors with missing dose information).

non peer reviewed
